# Effect of blood collection tube containing protease inhibitors on the pre-analytical stability of Alzheimer’s disease plasma biomarkers

**DOI:** 10.1101/2024.03.05.24303504

**Authors:** Yijun Chen, Xuemei Zeng, Jihui Lee, Anuradha Sehrawat, Tara K. Lafferty, James J. Boslett, William E. Klunk, Tharick A. Pascoal, Victor L. Villemagne, Annie D. Cohen, Oscar Lopez, Nathan A. Yates, Thomas K. Karikari

## Abstract

**INTRODUCTION:** The reliability of plasma Alzheimer’s disease (AD) biomarkers can be compromised by protease-induced degradation. This limits the feasibility of conducting plasma biomarker studies in environments that lack the capacity for immediate processing and appropriate storage of blood samples. We hypothesized that blood collection tube supplementation with protease inhibitors can improve the stability of plasma biomarkers at room temperatures (RT). This study conducted a comparative analysis of blood biomarker stability in traditional ethylenediaminetetraacetic acid (EDTA) tubes versus BD™ P100 collection tubes, the latter being coated with a protease inhibitor cocktail. The stability of six plasma AD biomarkers was evaluated over time under RT conditions.

**METHODS:** We evaluated three experimental approaches. In Approach 1, pooled plasma samples underwent storage at RT for up to 96 hours. In Approach 2, plasma samples isolated upfront from whole blood collected into EDTA or P100 tubes were stored at RT for 0h or 24h before biomarker measurements. In Approach 3, whole blood samples were collected into paired EDTA or P100 tubes, followed by storage at RT for 0h or 24h before isolating the plasma for analyses. Biomarkers were measured with Single Molecule Array (Simoa) and immunoprecipitation-mass spectrometry (IP-MS) assays.

**RESULTS:** Both the IP-MS and Simoa methods revealed that the use of P100 tubes significantly improved the stability of Aβ42 and Aβ40 across all approaches. Additionally, the Aβ42/Aβ40 ratio levels were significantly stabilized only in the IP-MS assay in Approach 3. No significant differences were observed in the levels of plasma p-tau181, GFAP, and NfL for samples collected using either tube type in any of the approaches.

**CONCLUSION:** Supplementation of blood collection tubes with protease inhibitors could reduce the protease-induced degradation of plasma Aβ42 and Aβ40, and the Aβ ratio for IP-MS assay. This has crucial implications for preanalytical procedures, particularly in resource-limited settings.

## 1. INTRODUCTION

Blood-based biomarkers of Alzheimer’s disease (AD) exhibit robust diagnostic and prognostic capabilities for the detection of disease-related changes across the disease continuum, including in asymptomatic individuals without cognitive impairment (Karikari *et al*. 2022; Teunissen *et al*. 2022; Gonzalez-Ortiz *et al*. 2023b; Hansson *et al*. 2022). This accumulating body of evidence forms the basis of calls for the clinical integration of blood biomarkers to enhance patient diagnosis and management (Karikari *et al*. 2022; Hansson *et al*. 2022). Furthermore, blood biomarkers are pivotal for clinical trials, proving instrumental for screening populations and identifying individuals with subtle or emerging biological abnormalities, thus facilitating their inclusion in long-term monitoring and therapeutic research (Hansson *et al*. 2022; Teunissen *et al*. 2022; Karikari *et al*. 2022; Gonzalez-Ortiz *et al*. 2023b). These developments underscore the substantial promise of blood-based diagnostics for neurodegenerative diseases.

Blood biomarkers for the amyloid beta (Aβ)42/40 ratio and phosphorylated tau (including p-tau181, p-tau217 and p-tau231) have demonstrated high potential to respectively detect brain Aβ plaques and tau neurofibrillary tangles, the principal pathological features of AD pathophysiology (Karikari *et al*. 2022; Gonzalez-Ortiz *et al*. 2023b; Balogun *et al*. 2023; Chong *et al*. 2021; Bellaver *et al*. 2023; Leuzy *et al*. 2022; Karikari *et al*. 2020; Gonzalez-Ortiz *et al*. 2024; Ferreira *et al*. 2023). In addition, plasma neurofilament light (NfL) is validated to be a good, albeit non-specific, biomarker for neurodegeneration/axonal injury while plasma glial fibrillary acidic protein (GFAP) is an indicator of astroglial activation often associated with brain Aβ plaques (Abdelhak *et al*. 2022; Ashton *et al*. 2021a; Benedet *et al*. 2021; Chatterjee *et al*. 2023). Whilst plasma Aβ42/40 and p-tau181 tend to become abnormal specifically in older adults with biological evidence of AD pathology when compared with those with other neurodegenerative diseases (Karikari *et al*. 2020; Ashton *et al*. 2021b; Janelidze *et al*. 2021a; Merluzzi *et al*. 2018), plasma NfL and GFAP have shown utility in AD [6, 7] and in other neurodegenerative diseases such as frontotemporal dementia (Ashton *et al*. 2021a; Zhu *et al*. 2021; Heller *et al*. 2020).

Widespread adoption of blood biomarkers is anticipated due to the ease, cost-effectiveness, and accessibility of blood-based sampling and analysis. To achieve this vision, preanalytical procedures must be adapted for use in studies conducted outside of well-equipped medical facilities (Karikari *et al*. 2022; Teunissen *et al*. 2022; Yiangou *et al*. 2021; Hansson *et al*. 2022). Current protocols emphasize immediate temperature-controlled storage of whole blood, followed by processing into plasma within 1-2 hours, beginning with centrifugation which requires reliable electricity supply (Karikari *et al*. 2022; Janelidze *et al*. 2022). These prerequisites are essential because plasma biomarkers, such as Aβ-based markers, are inherently unstable when stored at room temperature. However, meeting these conditions can be challenging in various research settings.

Proteolytic degradation is a primary factor contributing to the instability of blood biomarkers (Kurz *et al*. 2023; Yang *et al*. 2018; Saido & Leissring 2012). Previous studies demonstrated that treating plasma with protease inhibitors can attenuate the time-dependent reduction in Aβ42 levels and enhance the diagnostic accuracy of the Aβ42/Aβ40 ratio (Park *et al*. 2017). Meanwhile, several publications have suggested that using the BD™ P100 tubes, coated with a specialized protease inhibitor cocktail, can improve the stability of biomarker peptides in plasma during room temperature storage (Yi *et al*. 2008; Aguilar-Mahecha *et al*. 2012; Debunne *et al*. 2020). However, the utility of P100 tube for plasma AD biomarkers remains unknown. This study therefore aimed to evaluate the effectiveness of the P100 tube in stabilizing plasma AD biomarkers.

We first compared the room-temperature stability of various AD biomarkers in pooled plasma samples collected using either P100 tubes or conventional EDTA tubes (Fig. 1A). Subsequently, we expanded our investigation to encompass two distinct protocols, mimicking real-world scenarios where immediate centrifugation (Fig. 1B) or freezing (Fig. 1C) might not be practical. Our research findings revealed substantial time-dependent alterations in the concentrations of specific plasma biomarkers when whole blood or plasma is subjected to room temperature storage. Importantly, the use of the P100 tubes was observed to improve the preanalytical stability of these biomarkers. The assessed AD biomarkers included Aβ42, Aβ40, Aβ42/Aβ40 (evaluated through both IP-MS and Quanterix immunoassay), p-tau181, NfL, and GFAP (analyzed via Quanterix immunoassay).

**Figure 1.**
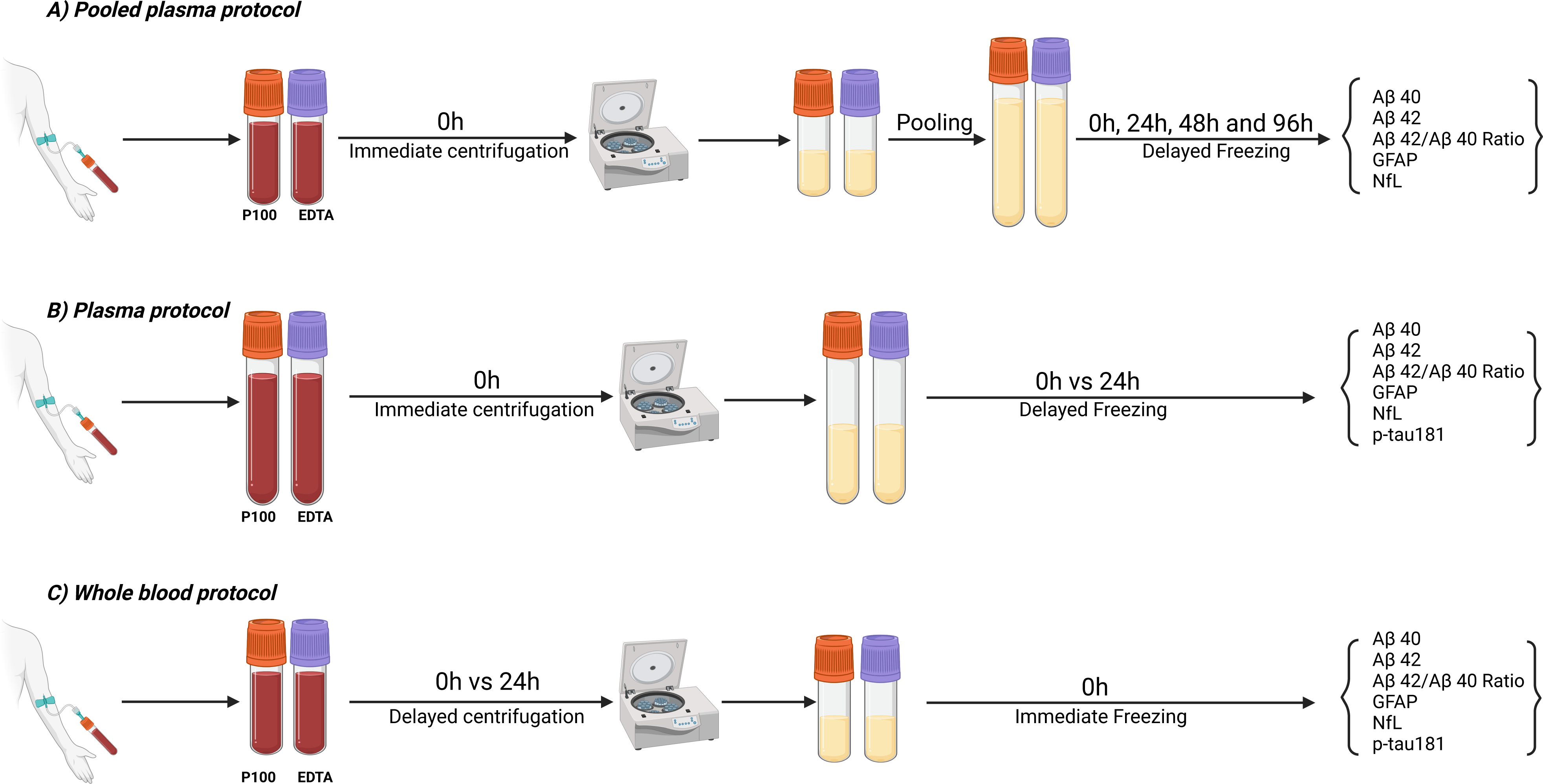
Schematic illustration of the experimental design. The *Pooled plasma protocol* (A) collection of whole blood samples from different patients into P100 and EDTA tubes individually, followed by pooling and storage at room temperature up to 96 hours. The *Plasma protocol* (B) involved immediate centrifugation of whole blood samples collected separately into P100 and EDTA tubes following standard procedures, followed by storage of the resulting plasma samples at room temperature for 24 hours. The *Whole blood* protocol (C), whole blood samples from the same blood draw were collected separately into P100 and EDTA tubes and stored directly at room temperature for 24 hours before obtaining the plasma by centrifugation and immediately freezing for subsequent analysis.

## 2. MATERIALS AND METHODS

### 2.1 Participants

We included participants enrolled in the University of Pittsburgh Alzheimer’s Disease Research Center (ADRC) cohort, who were undergoing annual visits as part of their regular clinical assessments which focused on diagnosis, neuroimaging, biomarker and neuropsychological testing. The present investigation was a prospective, blinded sub-study where participants were included according to their order of clinical attendance and informed consent to participate in the study including to have an extra volume of blood drawn. Participant information was unavailable to the study scientists until all the experiments had been completed and the statistical analyses had begun. The ADRC study was approved by the University of Pittsburgh Institutional Review Board (MOD19110245-023).

### 2.2 Blood collection and processing procedures

We followed standard procedures employed by trained ADRC clinical staff to collect blood for each experiment individually, with an experienced nurse performing all blood draws by venipuncture. Blood collection was performed between 9:00 am and 2:00 pm. In addition to routine ADRC blood collection, we collected additional blood tubes specifically for this study as detailed below.

A pilot experiment using pooled plasma samples from five patients was initially performed to test the performance of the P100 versus EDTA tubes (Approach 1). Subsequently, we assessed two distinct preanalytical protocols to simulate various scenarios. The “plasma protocol” (Approach 2) simulated conditions featuring on-site centrifugation but no ultra-freezing of plasma. The “whole blood protocol” (Approach 3), on the other hand, simulated resource-constrained settings where immediate centrifugation or frozen storage of processed plasma is unattainable. In all experimental approaches, we evaluated the preanalytical stability of plasma biomarkers in samples collected in standard EDTA tubes and in the P100 tubes from the same blood draw. To mitigate potential impacts of biological variation, samples from the same blood draw were aliquoted into pairs of tubes, with one in the EDTA tube and the other in the P100 tube.

In all experiments, room temperature was defined as the ambient temperature in the research laboratory (approximately 20 °C).

#### 2.2.1 Pooled plasma samples

The pooled plasma samples were obtained by combining plasma samples from five participants. Individual blood tubes collected either with 10 ml Lavender top EDTA tubes (BD Biosciences # 366643) or 8.5 ml P100 tubes (BD Biosciences #366448) were inverted 8 to 10 times immediately after the blood draw, and then centrifuged at 2000 xg for 15 minutes at 4°C to separate the plasma from the cellular material. Subsequently, plasma samples from each patient were pooled according to the tube type. Pooled samples were then aliquoted into cryovials and stored at room temperature for 0, 24, 48, or 96 hours (Fig. 1A).

#### 2.2.2 Plasma protocol

One 10 ml Lavender top EDTA tube and one 8.5 ml P100 tube were used to collect whole blood from each participant. After blood draw, the tubes were immediately inverted 8 to 10 times and centrifuged at 2000 xg for 15 min at 4°C to separate the plasma. The resulting plasma samples from each tube type were aliquoted into cryovials and stored at room temperature for either 0 or 24 hours after venipuncture (Fig. 1B).

#### 2.2.3 Whole blood protocol

Two of each 2 ml Lavender top EDTA tubes (BD Biosciences #367841) and 2 ml P100 tubes (BD Biosciences #366422) were used to collect whole blood. Following the blood collection, each tube was inverted 8 to 10 times immediately and then stored at room temperature for either 0 or 24 hours. After the specified storage period, the blood collection tubes were centrifuged at 2000 xg for 15 minutes at 4°C to isolate the plasma which were then aliquoted into cryovials and promptly frozen at -80°C until analysis (Fig. 1C).

### 2.3 Plasma biomarker measurements

The biomarker measurements were performed at the Biofluid Biomarker Laboratory, Department of Psychiatry, School of Medicine, University of Pittsburgh, PA, USA.

#### 2.3.1 Single molecule array (Simoa) assays

Simoa assays were performed on an HD-X instrument (Quanterix, Billerica, MA, USA). Plasma NfL, GFAP, Aβ42 and Aβ40 were measured with the Neurology 4-Plex E (#103670) and p-tau181 with the p-tau181 V2 Advantage kit (#103714) from Quanterix (Billerica, MA, USA). Quality control samples of three different concentrations were analyzed at the start and the end of each run to assess the reproducibility of each assay. The average within-run (p-tau181 = 4.1%, NfL = 9.2%, GFAP = 9.1%, Aβ42 = 5.1% and Aβ40 = 8.6%) and between-run (p-tau181 = 5.5%, NfL = 10.2%, GFAP = 13.5%, Aβ42 =6.9% and Aβ40 = 11.3%) coefficient of variation for quality control samples were below 15%.

#### 2.3.2. Immunoprecipitation with mass spectrometry (IP-MS) assay

The measurement of plasma Aβ40 and Aβ42 levels was conducted using an IP-MS method developed at the University of Pittsburgh based on the assay by Nakamura et al (Nakamura et al. 2018). Briefly, 250 μl of binding buffer (100 mM Tris-HCl pH 7.4 (Sigma #T2788-1L), 300 mM NaCl (Sigma #S7653-250G), 0.2% w/v n-dodecyl-β-D-maltoside (DDM; Sigma #D4641-1G), 0.2% w/v n-nonyl-β-D-thiomaltoside (Anatrace #148565-55-3)) containing 10 pM stable isotope labeled Aβ38 (Anaspec #AS-65220) was added to an 1.5 ml Eppendorf Protein LoBind tube (ThermoFisher #13-698-794), followed by the addition of 250 μl plasma sample. The samples were immunoprecipitated with 10 μl 50 mg/ml Dynabeads M-270 Epoxy (ThermoFisher #14301) coupled with 6E10 Aβ antibody (BioLegend #803003) for 1 hour at 4°C with rotation. After immunoprecipitation, the supernatant was discarded, and the beads were washed two times with 0.5 ml of cold PBS (Gibco #2537136). The washed beads were then transferred to a fresh Eppendorf tube and eluted with 25 μl of glycine elution buffer (50 mM glycine (pH 2.8, Sigma #G2879-100G), 0.1% DDM). The eluates were collected and transferred to fresh tubes containing 0.5 ml of binding buffer for a second round of immunoprecipitation. Thereafter, the beads were washed twice with 0.5 ml of cold HPLC-grade H2O (Fisher #7732-18-5) and then transferred to a fresh Eppendorf Protein LoBind Tube using 200 μl HPLC H2O. After removing the liquid through vacuum aspiration, Aβ peptides bound to the beads were eluted using 6 μl of 70% acetonitrile (Fisher #75-05-8) /5mM HCl (Fisher #7647-01-0). The eluate was spotted four times with 0.75 ul each onto MALDI target plate spots that were pre-spotted with 0.75 μl 2 mg/ml α-cyano-4-hydroxycinnamic acid matrix (Bruker #8201344). The MALDI target plate was air dried and then loaded into a benchtop time-of-flight mass spectrometer, Microflex LT (Bruker Daltonics) equipped with a 337 nm nitrogen laser to acquire mass spectra. The Microflex LT was operated in linear mode with a pulsed positive ion extraction setting. A peptide calibration mix (Bruker #8222570, #8206355) was used for the external mass calibration. One spectrum was acquired for each spot by combining ion signals from 2,500 laser shots, resulting in four spectra per sample. The acquired mass spectra were processed using ClinPro Tools Software (v2.1, Bruker Daltonics) for mass to charge alignment, peak detection, and peak area calculation.

The assay performance over different days was monitored using pooled quality control plasma samples. The inter-assay coefficients of variation for Aβ40 and Aβ42 were 10.4% and 13.6% respectively, while the intra-assay coefficients of variation were 13.9% for Aβ40 and 14.7% for Aβ42.

### 2.4 Statistical analyses

For demographic characteristics, continuous variables were summarized using median and interquartile range while categorical variables were reported as numbers and percentages. Kruskal-Wallis and Fisher’s exact tests were used to examine the difference across cohorts for continuous and categorical variables, respectively. Percent differences were calculated between all available biomarker measurements from individual samples at 24 hours and 0 hours, and these differences were compared between P100 and EDTA tubes using the Wilcoxon signed-rank test. A two-sided p-value less than 0.05 was considered statistically significant. All the analyses were conducted using R statistical software version 4.2.1 (R Foundation for Statistical Computing, Vienna, Austria; http://www.r-project.org/).

## 3. RESULTS

### 3.1 Participant characteristics

The main assessments included two cohorts, namely the “plasma cohort” and the “whole blood cohort” for evaluating the second and third approaches respectively (Fig. 1B, C). We included a total of 47 individuals, with 29 and 17 participants respectively. It is worth noting that one participant was included in both the plasma and whole blood protocols in the IP-MS assay, and, unfortunately, one sample failed in all assays. For the entire sample set, the median age was 76 years (Interquartile range [IQR]: 71.0 – 80.5 years), and there were 23 (48.9%) females. Twenty-three (48.9%) participants were *APOE* ε4 carriers, and 20 (42.6%) received a clinical diagnosis of AD according to published methods (Lopez *et al*. 2000a; Lopez *et al*. 2000b), but without biomarker assessments. In terms of cognitive performance, the median Mini-Mental State Examination (MMSE) and Montreal Cognitive Assessment (MoCA) scores were 25 (21.0 – 29.0), and 22 (17.0 – 26.5) respectively. Regarding Clinical Dementia Rating (CDR), 17 participants (36.2%) had a score of “mild or moderate.”

The demographic characteristics significantly varied solely in age between the plasma and whole blood cohorts used in the Simoa 4-plex (Aβ40, Aβ42, GFAP and NfL; p-value =0.041). Detailed information is shown in Table 1.

**Table 1.** The demographic of the patient samples.

|  |  | Gender<br>(Female) | Age | APOE ε4<br>genotype | Alzheimer's<br>Diagnosis<br>Positive | MMSE | MoCA | CDR |  |  |
| --- | --- | --- | --- | --- | --- | --- | --- | --- | --- | --- |
|  |  |  |  |  |  |  |  | Absent | Questionable | Mild or<br>Moderate |
| Total (N = 47) |  | 23 (48.9%) | 76 (71 - 80.5) | 23 (48.9%) | 20 (42.6%) | 25 (21 - 29) | 22 (17 - 26.5) | 10 (21.3%) | 20 (42.6%) | 17 (36.2%) |
| IP-MS<br>Aβ40<br>and<br>Aβ42§ | Plasma (N = 26) | 11 (42.3%) | 75 (69 - 77) | 11 (42.3%) | 11 (42.3%) | 27 (21.2 - 29) | 22 (16.2 - 27) | 5 (19.2%) | 10 (38.5%) | 11 (42.3%) |
|  | Whole blood (N = 15) | 9 (60%) | 79 (73 - 82) | 9 (60%) | 5 (33.3%) | 25 (21 - 29) | 23 (19 - 25.5) | 4 (26.7%) | 6 (40%) | 5 (33.3%) |
|  | p-value* | 0.341 | 0.061 | 0.341 | 0.742 | 0.576 | 0.968 |  | 0.915 |  |
| Simoa<br>Aβ40,<br>Aβ42,<br>GFAP,<br>NfL | Plasma (N = 21) | 9 (42.9%) | 75 (66 - 78) | 8 (38.1%) | 9 (42.9%) | 27 (22 - 30) | 22 (17 - 27) | 4 (19%) | 9 (42.9%) | 8 (38.1%) |
|  | Whole blood (N = 13) | 7 (53.8%) | 80 (73 - 85) | 7 (53.8%) | 5 (38.5%) | 24 (21 - 29) | 23 (17 - 25) | 5 (38.5%) | 4 (30.8%) | 4 (30.8%) |
|  | p-value* | 0.725 | **0.041** | 0.484 | 1.000 | 0.485 | 0.558 |  | 0.504 |  |
| Simoa<br>p-tau181 | Plasma (N = 17) | 10 (58.8%) | 75 (69 - 77) | 8 (47.1%) | 9 (52.9%) | 28 (22 - 30) | 22 (18 - 27) | 4 (23.5%) | 7 (41.2%) | 6 (35.3%) |
|  | Whole blood (N = 16) | 10 (62.5%) | 79.5 (73 - 83.5) | 9 (56.2%) | 5 (31.2%) | 25.5 (21 - 29) | 23 (19 - 25.2) | 5 (31.2%) | 6 (37.5%) | 5 (31.2%) |
|  | p-value* | 1.000 | 0.066 | 0.732 | 0.296 | 0.353 | 0.691 |  | 1.000 |  |

Median and interquartile range (IQR) are reported for continuous variables. Frequencies and percentages are shown for categorical variables.
§ One participant was included in both plasma and whole blood cohorts and thus accounted for reporting summary statistics in each cohort. \*P-values were calculated using Kruskal-Wallis test for a continuous variable and Fisher's exact test for a categorical variable, respectively.
Abbreviations: Aβ, amyloid beta; APOE, apolipoprotein E; A.U., absorbance unit; CDR, Clinical Dementia Rating; EDTA, ethylenediaminetetraacetic acid; GFAP, glial fibrillary acidic protein; IP-MS, immunoprecipitation-mass spectrometry; Simoa, single molecule array; MMSE, Mini Mental State Examination; MoCA, Montreal Cognitive Assessment; NfL, neurofilament light chain; p-tau181, tau phosphorylated at threonine-1

### 3.2 Time-dependent changes in pooled plasma samples collected into EDTA vs. P100 tubes

In the pilot experiment, we investigated the stability of plasma biomarkers using pooled plasma samples. The results indicated that plasma samples collected using EDTA tubes showed a sharp time-dependent decline in the concentrations of Aβ peptides (Aβ40 and Aβ42) over time, whereas samples collected using P100 tubes exhibited a slower decline (Fig. 2). In addition, the Aβ42/Aβ40 ratio demonstrated substantial changes over time according to the IP-MS results in the EDTA samples, whereas the results for P100 samples showed less pronounced changes (Fig. 2). In contrast, the concentrations of GFAP, NfL, and p-tau181 remained stable across multiple time points, irrespective of whether EDTA or P100 tubes were utilized (Supplementary. Fig. 1). A detailed summary of the time-dependent changes in each marker can be found below (Table 2). Due to the small sample size and low power of this initial experiment, no statistical analysis was performed in this experiment.

**Figure 2.**
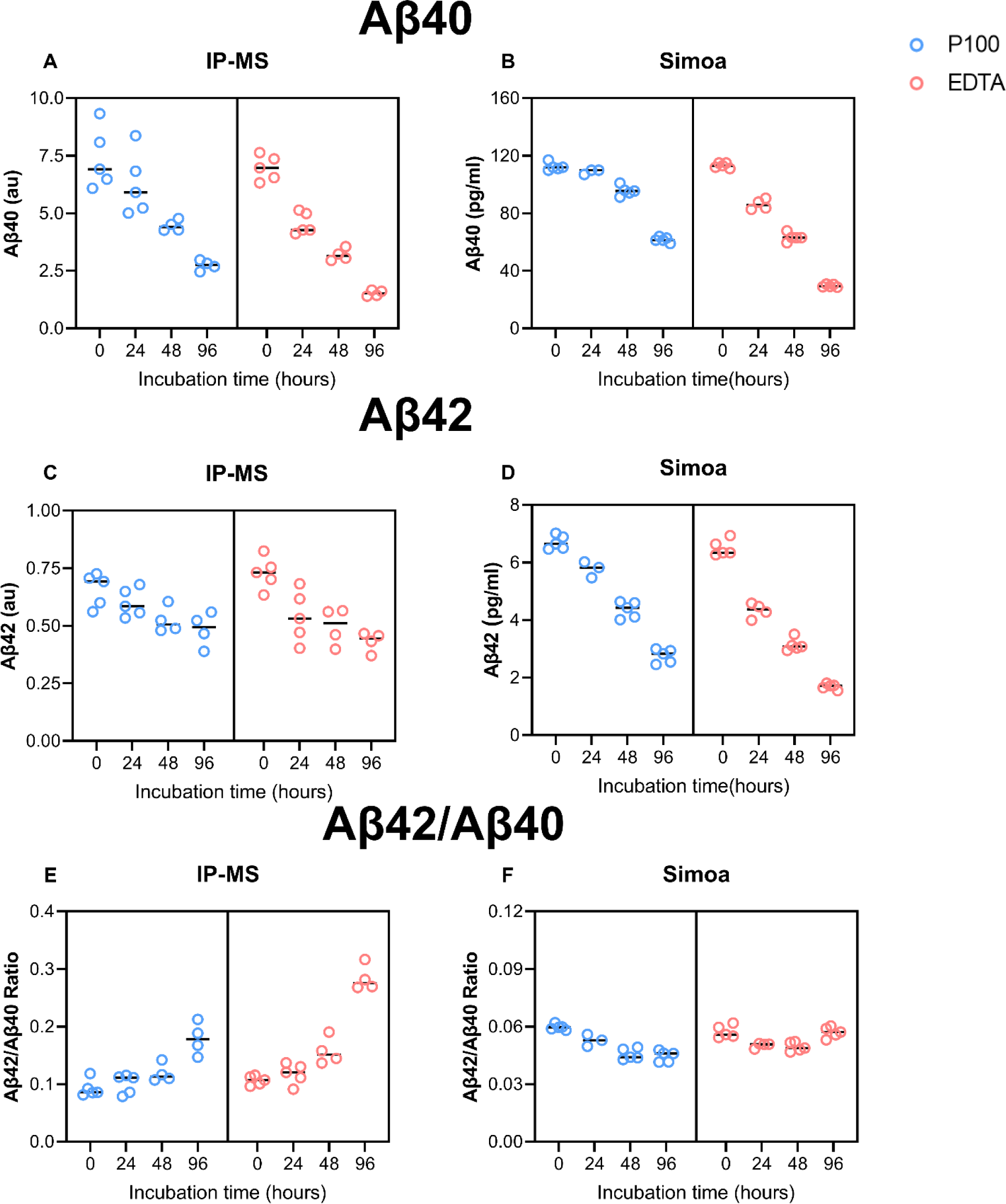
Time-dependent changes in pooled plasma Aβ40 and Aβ42 concentrations and the Aβ42/Aβ40 ratio over 96 hours of storage at room temperature in the pooled plasma experiment. This figure shows the time-dependent changes in plasma Aβ40 (A, B) and Aβ42 (C, D) concentrations and the Aβ42/Aβ40 ratio (E, F) at four time points: 0, 24, 48, and 96 hours. These alterations are depicted for pooled samples that were initially collected in both P100 and EDTA tubes. The results are presented for both the IP-MS and the Simoa assays. The data points shown are up to five technical replicates for the pooled samples at the given timepoints. The median concentration is depicted as the central horizontal bar.

**Table 2.**
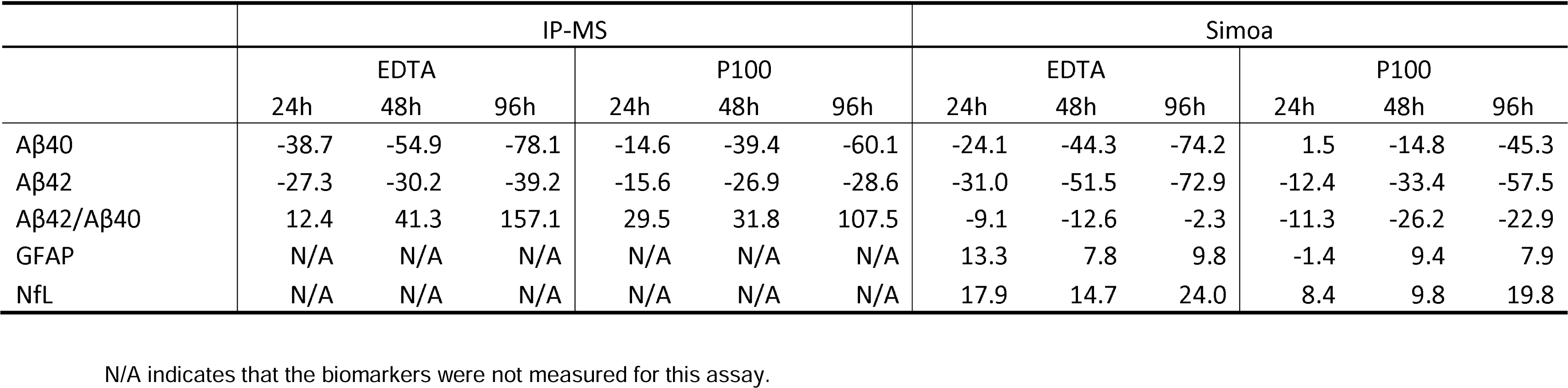
Time-dependent changes in pooled plasma samples (%)

#### Aβ40

There was a time-dependent decline in Aβ40 abundance when samples were stored at room temperature. For samples collected in EDTA tubes, IP-MS Aβ40 showed median percent changes of -38.7%, -54.9%, and -78.1% after 24, 48 and 96 hours respectively (Fig. 2A). The Simoa EDTA results closely reproduced these findings, with a median percent change of -24.1% after 24 hours, -44.3% after 48 hours, and -74.2% after 96 hours (Fig. 2B). In contrast, the percent change in Aβ40 for samples collected using the P100 tubes exhibited a less steep decline. The IP-MS results showed median percent changes of -14.6%, -39.4% and -60.1% after 24, 48 and 96 hours respectively (Fig. 2A). The Simoa results recorded median percent changes of -1.5%, -14.8% and -45.3% after 24, 48 and 96 hours respectively (Fig. 2B).

#### Aβ42

The Aβ42 results were similar to those for Aβ40. For samples collected in EDTA tubes, the IP-MS results indicated a -27.3% change in median after 24 hours of storage, -30.2% after 48 hours, and -39.2% after 96 hours (Fig. 2C). For Simoa, there were -31.0%, -51.5% and -72.9% changes in Aβ42 concentration after 24, 48 and 96 hours respectively (Fig. 2D). On the other hand, the percent change for samples collected using P100 tubes recorded less signal decline. The IP-MS results showed a -15.6% change in median after 24 hours of storage, -26.9% after 48 hours, and -28.6% after 96 hours (Fig. 2C). For the Simoa results, the percent changes were -12.4%, -33.4% and -57.5% after 24, 48 and 96 hours respectively (Fig. 2D).

#### Aβ42/ Aβ40 ratio

For samples collected in EDTA tubes, the IP-MS results showed a 12.4% change in levels after 24 hours, 41.3% after 48 hours, and 157.1% after 96 hours (Fig. 2E). For the Simoa assay, the ratio changes were -9.1%, -12.6%, and -2.3% after 24, 48 and 96 hours respectively; however, these changes were not significant (Fig. 2F). The percent change for samples collected using P100 tubes was 29.5%, 31.8%, and 107.5% after 24, 48 and 96 hours respectively (Fig. 2E). In Simoa, the percent changes of the ratio were -11.3%, -26.2%, and -22.9% after 24, 48 and 96 hours respectively (Fig. 2F).

#### GFAP and NfL

For GFAP and NfL, the changes in concentrations over multiple time points were less. Specifically, the results for GFAP in EDTA tubes demonstrated a 13.3%, 7.8% and 9.8% change after 24, 48 and 96 hours (Supplementary Fig. 1A, Supplementary Fig. 1B). In contrast, for the P100 tubes, the results were -1.4%, 9.4%, and 7.9% after 24, 48 and 96 hours respectively, indicating relatively stable concentrations. Regarding NfL, the results showed a 17.9% change after 24 hours, 14.7% after 48 hours, and 24.0% after 96 hours for EDTA tubes. Conversely, for P100 tubes, the results were 8.4%, 9.8%, and 19.8% after 24, 48 and 96 hours respectively, demonstrating a similar trend of relatively stable concentrations (Supplementary Fig. 1A-B).

### 3.3 Effect of using P100 blood tubes on plasma biomarker stability in samples experiencing delayed freezing

In the plasma protocol, for EDTA, the plasma biomarker results showed that the Aβ peptides (Aβ40 and Aβ42) were the most affected while those collected using the P100 tubes showed significant improvements in addressing this loss of stability (Fig. 3). However, since both peptides were affected, the change in the Aβ42/Aβ40 ratio was not significant, just as the concentrations of GFAP, NfL, and p-tau181 were not significantly different between the 0-hour and 24-hour time points, irrespective of whether EDTA or P100 tubes were employed (Fig.3, Supplementary Fig. 2). A detailed description of the results of each marker can be found below (Table 3).

**Figure 3.**
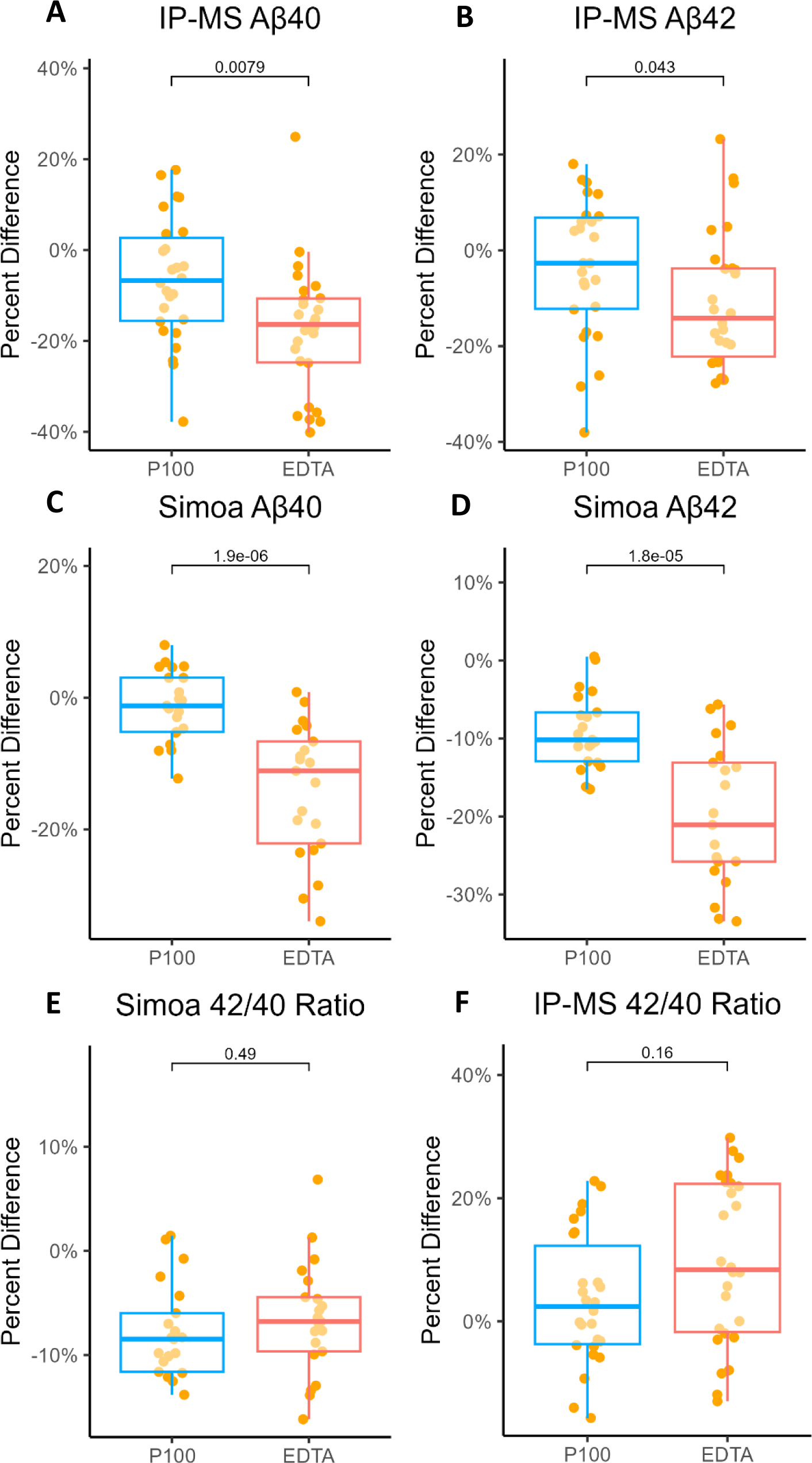
Changes in plasma Aβ40 and Aβ42 concentrations and the Aβ42/Aβ40 ratio over 24 hours of storage at room temperature according to the plasma protocol. This figure illustrates the alterations in plasma Aβ40 and Aβ42 concentrations and the Aβ42/Aβ40 ratio over a 24-hour period for paired samples initially collected in both P100 and EDTA tubes. The results are presented for both the IP-MS and Simoa assays using samples obtained through the plasma protocol. In the boxplots, the median concentration is depicted as the central horizontal bar, flanked by the 25th and 75th percentile values. Outliers are represented as individual data points. Statistical significance was determined using the Wilcoxon signed-rank test.

**Table 3.** Biomarkers abundance change (%)

| Plasma protocol |  |  |  |  |  |  |
| --- | --- | --- | --- | --- | --- | --- |
|  | IP-MS |  |  | Simoa |  |  |
|  | EDTA | P100 | p-value | EDTA | P100 | p-value |
| Aβ40 | -16.4 | -6.7 | **7.9E-03** | -11.1 | -1.2 | **1.9E-06** |
| Aβ42 | -14.2 | -2.7 | **0.043** | -21.1 | -10.2 | **1.8E-05** |
| Aβ42/Aβ40 | 8.4 | 2.4 | 0.16 | -6.8 | -8.5 | 0.49 |
| GFAP | N/A | N/A | N/A | 12.1 | 2.2 | 0.050 |
| NfL | N/A | N/A | N/A | 8.7 | 6.6 | 0.050 |
| p-tau181 | N/A | N/A | N/A | 1.6 | 1.4 | 0.97 |

| Whole blood protocol |  |  |  |  |  |  |
| --- | --- | --- | --- | --- | --- | --- |
|  | IP-MS |  |  | Simoa |  |  |
|  | EDTA | P100 | p-value | EDTA | P100 | p-value |
| Aβ40 | -34.8 | -16.7 | **8.4E-03** | -48.3 | -25.6 | **2.4E-04** |
| Aβ42 | -23.0 | -17.9 | 0.30 | -46.0 | -31.3 | **2.4E-04** |
| Aβ42/Aβ40 | 11.4 | 6.0 | **0.018** | -3.2 | -8.1 | 0.74 |
| GFAP | N/A | N/A | N/A | 0.4 | 6.0 | 0.22 |
| NfL | N/A | N/A | N/A | -1.9 | 3.4 | 0.19 |
| p-tau181 | N/A | N/A | N/A | -0.2 | -0.3 | 0.38 |

N/A indicates that the biomarkers were not measured for this assay.
P-values were calculated using Wilcoxon test.

#### Aβ40

In the case of samples collected in EDTA tubes, we observed a median percent change of -16.4% in the IP-MS Aβ40 signal when comparing samples stored for 24 hours versus 0 hours at room temperature (Fig. 3A). Similar results were recorded for Simoa Aβ40, with a median percent change of -11.1% at 24 hours (Fig. 3C). In contrast, plasma sample aliquots processed from the P100 tubes exhibited significantly greater stability, with median signal changes of -6.7% and - 1.2% for the IP-MS and Simoa Aβ40 assays (Fig. 3A, Fig. 3C). Statistical analysis using Wilcoxon tests revealed significant differences (p-value = 0.0079 for IPMS, p-value = 1.9e-6 for Simoa) in the percent difference of Aβ40 levels between samples collected using EDTA tubes and P100, as measured by either assay.

#### Aβ42

In EDTA samples stored at room temperature for 24 hours, there was a -14.2% median percent change in IP-MS Aβ42 signals and a -21.1% change in Simoa Aβ42 signals compared to the 0-hour measurements (Fig. 3B, Fig. 3D). However, plasma samples collected from the P100 tubes exhibited greater stability, with only -2.7% change for the IP-MS assay and -10.2% for Simoa (Fig. 3B, Fig. 3D). Statistical analysis using Wilcoxon tests indicated significant differences (p-value =0.043 for IPMS, p-value =1.8e-5 for Simoa) in the percent difference of Aβ42 levels between samples collected using EDTA and P100 tubes, as measured by either assay.

#### Aβ42/ Aβ40 ratio

Since both Aβ peptides showed decreased levels, the Aβ42/Aβ40 ratio did not exhibit a significant difference in the percent change between the two tube types over 24 hours storage. For samples collected using EDTA tubes, the median percent difference over 24 hours of room temperature storage was 8.4% for IP-MS and -6.8% for the Simoa assay (Fig. 3E, Fig. 3F). The P100 tube results showed a median percent difference of 2.4% and -8.5% according to IP-MS and Simoa assays, respectively (Fig. 3E, Fig. 3F).

#### GFAP, NfL and p-tau181

The percent difference in signal between the two timepoints for the samples collected using EDTA tubes were 12.1% for GFAP, 8.7% for NfL, and 1.6% for p-tau181, which were not statistically different from 2.2%, 6.6% and 1.4% for samples collected using P100 (Supplementary Fig. 2).

### 3.4 Storage of whole blood in P100 tubes partially addresses effects of delayed centrifugation

In the whole blood protocol, the levels of Aβ42 and Aβ40 in EDTA samples both exhibited decreasing trends over time, as observed using both the IP-MS and Simoa methods. However, the use of P100 tubes mitigated this degradation. The Aβ42/Aβ40 ratio remained stable throughout the observation period, regardless of whether we used EDTA or P100 tubes, as shown by the Simoa results. However, the IP-MS results indicated changes over 24 hours for samples collected with EDTA tubes, while samples collected with P100 tubes remained stable (Fig. 4). On the other hand, the concentrations of GFAP, NfL, and p-tau181 remained consistent between the 0-hour and 24-hour time points, regardless of whether EDTA or P100 tubes were used (Supplementary Fig. 3). The results are summarized in Table 3.

**Figure 4.**
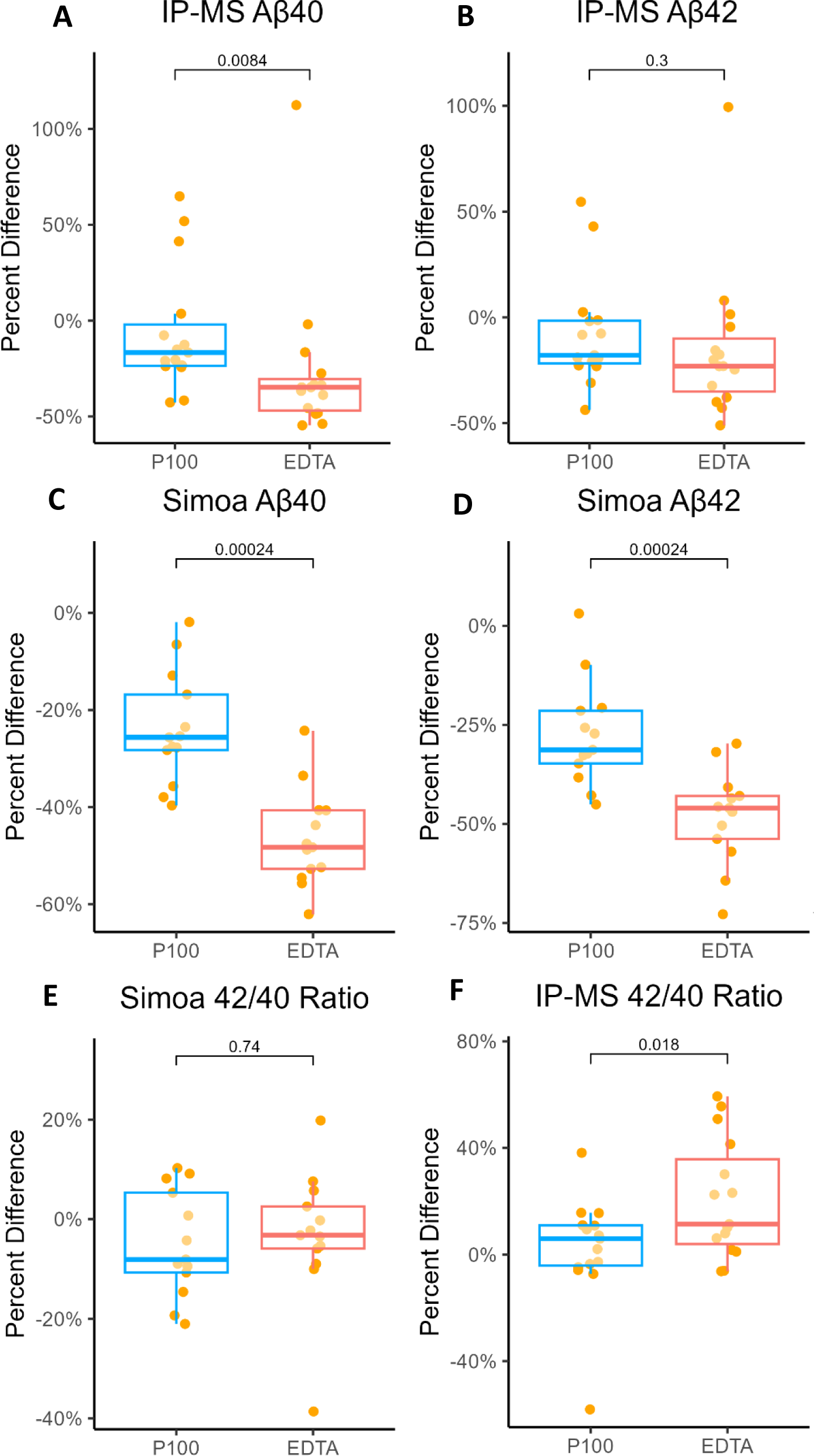
Changes in plasma Aβ40 and Aβ42 concentrations and the Aβ42/Aβ40 ratio over 24 hours of storage at room temperature according to the whole blood protocol. This figure illustrates the alterations in plasma Aβ40 and Aβ42 concentrations and the Aβ42/Aβ40 ratio over a 24-hour period for paired samples initially collected in both P100 and EDTA tubes. The results are presented for both the IP-MS and Simoa assays using samples obtained through the whole blood protocol. In the boxplots, the median concentration is depicted as the central horizontal bar, flanked by the 25th and 75th percentile values. Outliers are represented as individual data points. Statistical significance was determined using the Wilcoxon signed-rank test.

#### Aβ40

In the case of samples collected in EDTA tubes, we observed a substantial decrease in the median percent change of IP-MS Aβ40 signal by -34.8% when comparing samples stored for 24 hours versus those at 0 hours at room temperature (Fig. 4A). Similarly, the results from the Simoa Aβ40 assay displayed a notable -48.3% median signal change at 24 hours (Fig. 4C). In contrast, plasma sample aliquots processed from P100 tubes exhibited significantly greater stability, with median percent changes of only -16.7% and -25.6% between the initial 0-hour measurements and those taken at 24 hours for the IP-MS and Simoa Aβ40 assays respectively (Fig. 4A, Fig. 4C). Statistical analysis employing Wilcoxon tests underscored significant differences (p-value =0.0084 for IPMS, p-value =0.00024 for Simoa) in the percent difference of Aβ40 levels between samples collected using EDTA tubes and P100, as measured by either assay.

#### Aβ42

When assessed using IP-MS, the EDTA tubes exhibited a median percent difference of -23.0% versus -17.9% for P100 tubes (Fig. 4B). The Wilcoxon test indicated no significant difference between the two tube types for IP-MS data. However, a notable distinction emerged when considering Aβ42 levels based on the Simoa data results. For samples collected using EDTA tubes, the median percent difference over 24 hours of room temperature storage was a substantial -46.0% (Fig. 4D). In contrast, the results from P100 tubes showed a significantly (p-value =0.00024) smaller median percent difference of -31.3% (Fig. 4D).

#### Aβ42/ Aβ40 ratio

When assessed using IP-MS, there was a median percent difference of 11.4% for samples collected in EDTA tubes, whereas P100 tubes demonstrated better stability with a median percent difference of only 6.0% (Fig. 4F). Wilcoxon tests indicated a significant difference (p-value =0.018) between the two tube types for IP-MS data. In contrast, the Aβ42/Aβ40 ratio did not exhibit a significant difference in the percent change between the two tube types based on the Simoa data results. For samples collected using EDTA tubes, the median percent difference over 24 hours of room temperature storage was -3.2% and -8.1% for the P100 tubes (Fig. 4E).

#### GFAP, NfL and p-tau181

The concentrations of GFAP, NfL, and p-tau181 exhibited minimal change between the 0-hour and 24-hour time points. The percent change in signal for EDTA tubes was 0.4%, -1.9%, and - 0.2%, which did not show a statistically significant difference compared to 6.0%, 3.4%, and -0.3% for samples collected using the P100 tubes. (Supplementary Fig. 3). These findings showed no significant difference in the percent difference between the two tube types based on the Wilcoxon test.

## 4. DISCUSSION

Numerous studies have reported the vulnerability of plasma AD biomarkers, particularly Aβ40 and Aβ42, to preanalytical degradation when stored at room temperature for prolonged periods, emphasizing concerns about their stability during delayed centrifugation or freezing in resource-limited settings (Ashton *et al*. 2021c; Kurz *et al*. 2023; Sunde *et al*. 2023; Verberk *et al*. 2022). In this study, we aimed at addressing the challenge of stabilizing plasma AD biomarkers by investigating the effectiveness of P100 tubes coated with protease inhibitors. We began with pooled plasma samples, demonstrating the effectiveness of P100 tubes, and later evaluated individual samples using two distinct experimental approaches: plasma protocol and whole blood protocol. We observed improved stability of Aβ peptides in both settings when P100 tubes were used to collect blood instead of traditional EDTA tubes. Our analysis covered a wide panel of plasma AD biomarkers and examined their stability under real-world conditions mimicking resource-limited settings.

Significant enhancements in signal stability were evident, particularly for the most vulnerable markers, specifically Aβ40 and Aβ42, when utilizing P100 tubes for both assays. Moreover, the result of the IP-MS assay demonstrated notable improvements in the stability of Aβ42/Aβ40 ratio when utilizing P100 tubes. However, this result was only limited to the pooled plasma approach and whole blood protocol. Conversely, with the Simoa assay, the changes in the Aβ42/Aβ40 ratio were not as prominent as those observed for the individual peptides. This can be explained by the observation that both peptides exhibited decreases in the EDTA tubes and P100 tubes. As a result, these decreases canceled each other out, leading to a non-significant change in the Aβ42/Aβ40 ratio. We did not record significant changes in the stability of GFAP, NfL and p-tau181 in samples collected using P100 versus EDTA tubes. All three biomarkers remained stable at room temperature, which is consistent with previous results (Ashton *et al*. 2021c; Verberk *et al*. 2022; Altmann *et al*. 2021; Bowen *et al*. 2010). Since plasma biomarkers are often measured as a panel, utilizing P100 tubes has an added advantage when all the analytes examined herein are of interest.

More substantial enhancements in the preanalytical stability of the Aβ peptides were observed in the plasma samples compared with the whole blood samples. While the exact reason for this observation remains unclear, previous reports have suggested that protease inhibitors are more active in plasma than whole blood samples (Yi *et al*. 2015). Others have identified that hematological changes in whole blood that occur during storage induce hemolysis, releasing various enzymes including proteases from red blood cells (Sawant *et al*. 2007). These released proteases have the potential to exacerbate the degradation of the proteins/peptides of interest. In that case, the protease inhibitor may not perform as effectively as in the plasma samples.

Additionally, we observed some variations in results between the IP-MS and Simoa assays, particularly regarding Aβ42 levels and the Aβ42/40 ratio. For instance, the IP-MS assay, but not the Simoa assay, showed significant difference in the changes of Aβ42/Aβ40 ratio over time between samples collected using P100 tubes and EDTA tubes within the pooled plasma samples and the whole blood samples. These disparities can be attributed to the inherent differences of non-specific interference and sensitivity in the two assay platforms. The Simoa assay is an immunoassay that has enhanced sensitivity. However, like all immunoassays, it may be susceptible to non-specific interference. It has been reported by Thijssen et al. that besides Aβ1-42, the Quanterix Aβ42 assay also recognized Aβ2-42 and Aβ3-42, making it more of an Aβx-42 assay recognizing all peptides ending at Aβ42 without being specific to the N-terminal end of the peptide (Thijssen *et al*. 2021). In contrast, our IP-MS assay employs Aβ antibodies to enrich specifically for Aβ1-40 and Aβ1-42 peptides from plasma samples, followed by MS-based quantification. The IP-MS has lower sensitivity compared to the Simoa assay. However, it provides superior specificity by incorporating the mass-to-charges of Aβ peptides. In agreement, a head-to-head comparison study has suggested superior diagnostic performance for IP-MS based Aβ measurements over immunoassay-based measurement (Janelidze *et al*. 2021b).

Our results may prove useful in several resource limited scenarios. Firstly, many community- and population-based studies shifted to home visits for blood collection, notably during the COVID-19 pandemic. Moreover, many low- and middle-income countries grapple with unreliable electricity access, particularly for temperature-controlled storage and centrifugation. Lastly, immediate processing and storage of blood samples collected in the field from active-duty servicemen is often unattainable. Therefore, less resource-intensive preanalytical protocols applicable to these settings are needed to enable the power of blood biomarkers to help generalize access to diagnostics and screening tools for AD and related neurodegenerative diseases.

Our study has several notable strengths. Our primary focus remains to facilitate plasma biomarker collection and analyses in resource-limited settings while rigorously adhering to a standardized procedure for sample collection. All samples were obtained within a specific time frame to minimize potential external influences. The study assessed Aβ40 and Aβ42 through two separate assay modalities: immunoassay and mass spectrometry assay, providing comprehensive insights into Aβ processing. Compared with other approaches such as dry blood spot and finger-prick methods which lead to substantially different biomarker concentration values and increased sample processing workload that can also be a major source of preanalytical variability (e.g., reconstituting the filter paper in buffer), the P100 approach described herein enables more straightforward applications.

Limitations of this study include the following. Firstly, the sample size across cohorts was relatively small. Future evaluation utilizing a larger scale and ideally in real-world settings will be needed to confirm our findings. Additionally, due to commercial confidentiality reasons, the specific protease inhibitor information used in the P100 tube remains undisclosed. Additional research would be needed to pinpoint which specific protease inhibitor(s) contribute(s) to the stabilization of plasma Aβ peptides. Other novel blood biomarkers including plasma p-tau217, p-tau231, and brain-derived tau (Gonzalez-Ortiz *et al*. 2023c; Gonzalez-Ortiz *et al*. 2023a; Gonzalez-Ortiz *et al*. 2023d) were also not evaluated.

We have shown that the utilization of P100 tubes can help stabilize Aβ peptides in both plasma and whole blood samples, with exclusive improvement in the Aβ42/40 ratio observed in the IP-MS assay. This stabilization effect was observed without affecting the measurements of GFAP, NfL, and p-tau181 biomarkers. Our findings suggest that P100 tubes offer improvement in maintaining the stability of plasma Aβ peptides. After further optimization, this approach has the potential to be useful to enable the expansion of AD plasma biomarker analyses to resource-limited settings where the standard procedures of immediate temperature-controlled centrifugation of whole blood and ultra-freezing of the resulting plasma samples may not be feasible. This includes, but not limited to, community- and population-based studies where blood is collected during home visits, primary care centers in small towns and villages as well as health facilities in low- or middle-income countries without the resources to process and store blood following standard operating procedures.

## Data Availability

All data produced in the present study are available upon reasonable request to the authors

## Acknowledgements

The authors would like to express their sincere gratitude to all the participants who generously donated their blood for this research. Special thanks are extended to Arlene Malia from the University of Pittsburgh ADRC for her invaluable time and expertise in collecting blood samples. The authors also extend their gratitude to other members of the Karikari laboratory, including Carl Bertram, for their support and contributions. Furthermore, the authors would like to acknowledge Harris Bell-Temin for his valuable discussions and suggestions regarding the examination of the P100 blood collection system.

