## Supplementary material for "Effect of blood collection tube containing protease inhibitors on the pre-analytical stability of Alzheimer’s disease plasma biomarkers": Supplymental

**Supplementary Figure 1.**

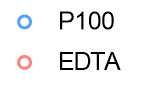

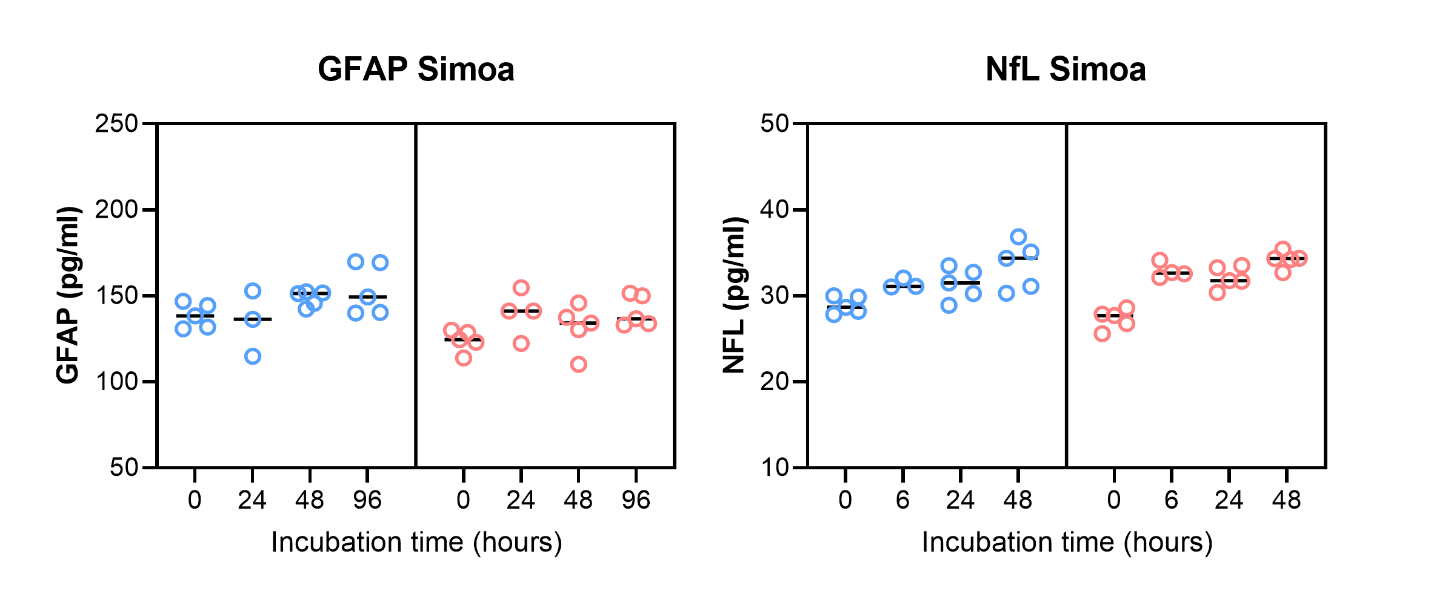

**B**

**A**

**Supplementary Figure 2.**

**A**

**B**

**C**

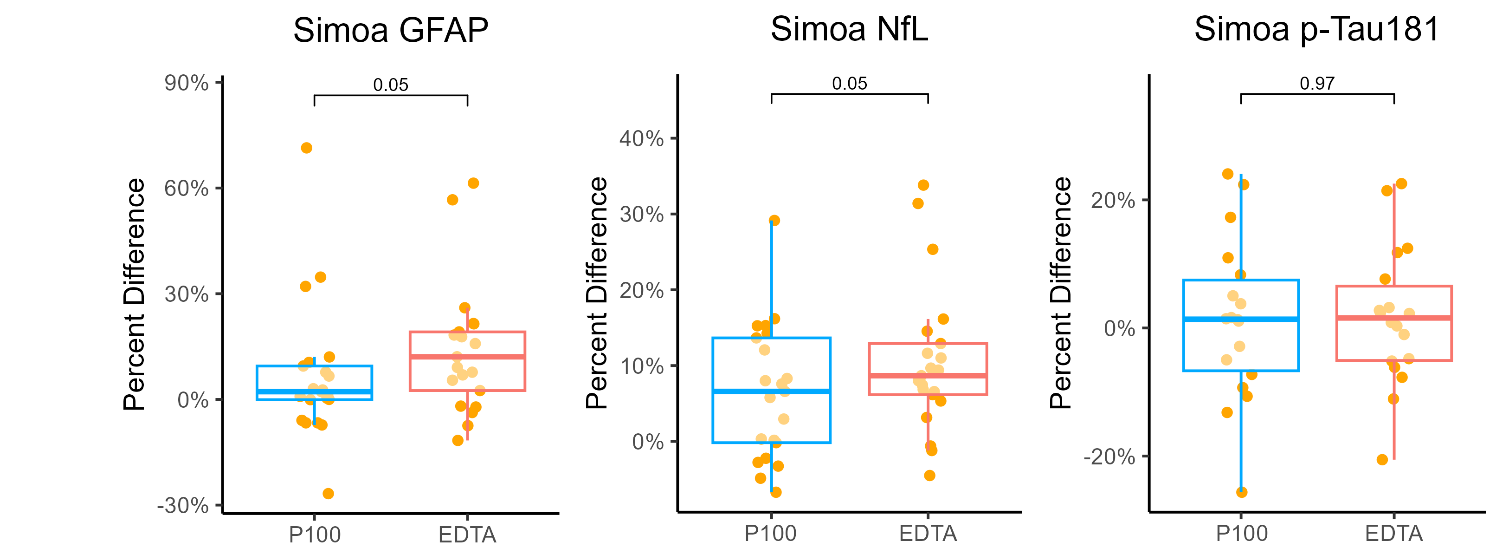

**Supplementary Figure 3.**

**C**

**B**

**A**

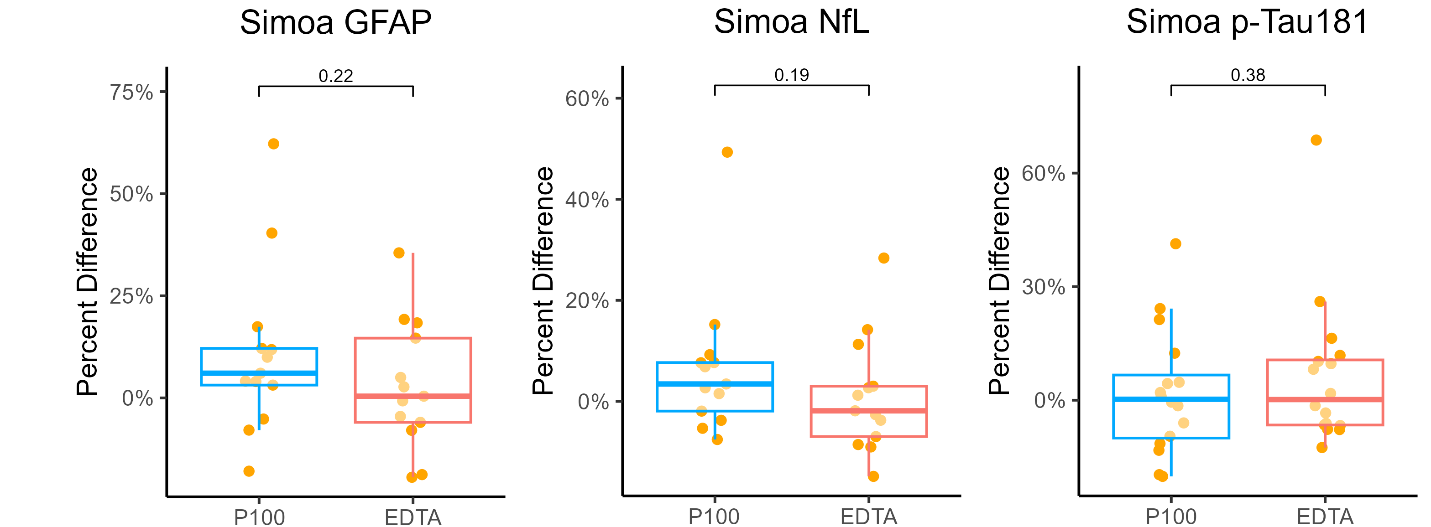

**Figure legends**

**Supplementary Figure 1. Time-dependent changes in the concentrations of GFAP and NfL in the pooled plasma sample experiment over 96 hours.** This figure shows the changes in plasma GFAP and NfL concentrations measured in the plasma samples obtained by pooling aliquots from different patients (see Figure 1) over four three-time intervals: 0, 24, 48, and 96 hours. These alterations are depicted for pooled samples that were initially collected in both P100 and EDTA tubes. The results are presented for Simoa assay. In the point plots, the median concentration is depicted as the central horizontal bar.

**Supplementary Figure 2. Changes in plasma GFAP, NfL and p-tau181 concentrations over 24 hours in the plasma protocol.** This figure illustrates the alterations in plasma GFAP, NfL and p-Tau181 over a 24-hour period for paired samples initially collected in both P100 and EDTA tubes. The results are presented for the Simoa assay using samples obtained through the plasma protocol. In the boxplots, the median concentration is depicted as the central horizontal bar, flanked by the 25^th^ and 75^th^ percentile values. Outliers are the individual data points outside of the box. Statistical significance was determined using the Wilcoxon signed-rank test.

**Supplementary Figure 3. Changes in plasma GFAP, NfL and p-Tau181 concentrations over 24 hours in the whole blood protocol.** This figure shows the alterations in plasma GFAP, NfL and p-Tau181 over a 24-hour period for paired samples initially collected in both P100 and EDTA tubes. The results are presented for the Simoa assay using samples obtained through the whole blood protocol. In the boxplots, the median concentration is depicted as the central horizontal bar, flanked by the 25th and 75th percentile values. Outliers are represented as individual data points. Statistical significance was determined using the Wilcoxon signed-rank test.
